# Rapid environmental monitoring, capture, and destruction activities of SARS-CoV-2 during the Covid-19 health emergency

**DOI:** 10.1101/2020.11.24.20237040

**Authors:** Roberto Marchetti, Martina Stella, Debjyoti Talukdar, Rosaria Erika Pileci

**Affiliations:** Laboratori Clodia Diagnostics & Services, Bolzano, Italy; Teerthanker Mahaveer University, Moradabad Campus, Uttar Pradesh, India; U-Earth Biotech Ltd, Milan, Italy

**Keywords:** biomonitoring, environmental monitoring, occupational surveillance, workplace exposures, disease, inhalation exposure, air purification

## Abstract

**Objectives:** SARS-CoV-2 pandemic is a health emergency for occupational healthcare workers at COVID19 hospital wards in Italy. The objective of the study was to investigate if U-Earth AIRcel bioreactors were effective in monitoring and improving air quality via detection, capture, and destruction of the SARS-CoV-2 virus, reducing the risk of transmission among healthcare workers.

**Methods:** U-Earth AIRcel bioreactors are a demonstrated effective biomonitoring system. We implemented a methodological approach wherein they were placed at various hospitals treating COVID-19 patients in Italy. The detection of the SARS-CoV-2 virus was achieved through rapid biomonitoring testing of the solutes from the AIRcel bioreactors via SARS-CoV-2 rapid test antigen and consecutive reverse transcription-polymerase chain reaction (RT-PCR) analysis with the multiplex platform (XABT) and the Real-Time PCR Rotor-Gene.

**Results:** The marked presence of the SARS-CoV-2 virus was found in multiple water samples via the detection of ORF1ab + N and/or E gene involved in gene expression and cellular signaling of the SARS-CoV virus. The AIRcel bioreactors were able to neutralize the virus effectively as traces of the viruses were no longer found in multiple solute samples after an overnight period.

**Conclusions:** Transmission of COVID-19 via bio-aerosols, transmitted by infected patients, remains a viable threat for health workers. AIRcel bioreactors allow for rapid biomonitoring testing for early virus detection within the environment, reducing the risk of exponential contagion exposure and maintaining good air quality without endangering health workers. This same protocol can also be extended to public spaces as a bio-monitoring tool for hotpots early detection.

**Key messages:** *What is already known about this subject?:*

- Transmission of SARS-CoV-2 virus via bio-aerosols is a threat to health care workers. Only few studies have conducted investigations on how to limit the spread of the virus via air purifiers.
- Existing studies show a higher risk to health care workers serving at COVID-19 wards with a higher risk of viral transmission.

*What are the new findings?:*

- In this study, SARS-CoV-2 virus traces were captured by U-Earth air purifier bioreactor units placed at several hospitals in Italy.
- AIRcel bioreactors achieved early detection of the SARS-CoV-2 virus within the environment via rapid biomonitoring testing.
- AIRcel bioreactors have proved effective in biomonitoring via the detection, capture, and destruction of SARS-CoV-2 virus through reverse transcription-polymerase chain reaction (RT-PCR) analysis with the multiplex platform (XABT) Multiple Real-Time PCR Rotor-Gene.

*How might this impact on policy or clinical practice in the foreseeable future?:*

- This study shows the need for effective surveillance and biomonitoring to contain the spread of the SARS-CoV-2 virus. AIRcel bioreactors, an effective occupational surveillance system, can reduce the transmission of the virus to health care workers serving COVID-19 infected patients at hospital wards.
- AIRcel bioreactors can also be used in public spaces and other settings, such as schools, to increase the speed of detection of the SARS-CoV-2 virus and improve control of the environment, thereby decreasing the exponential growth of the pandemic.

## INTRODUCTION

Coronaviruses are part of a large family of viruses that infect humans and animals, and are classified into 4 genera (α, β, γ, and δ). Human coronaviruses belong to α and β genera (1). The current outbreak of β-coronavirus disease, emerged in Wuhan in December 2019 (COVID-19) and named severe acute respiratory syndrome coronavirus 2 (SARS-CoV-2) has led to a pandemic (June 28^th^, 2020 WHO report), which, at the time of paper submission, is affecting more than 55.6 million people, with more than 1.3 million deaths in 231 countries (2–4). This pandemic outbreak, with regards to the ease and astonishing speed with which the virus spreads, has highlighted the necessity to implement preventive surveillance systems for viruses that are proactive and allow rapid isolation actions.

The significance of SARS-CoV-2 viral transmission via aerosols (airborne transmission) has been intensely discussed and it is now accepted to be among the pathways of viral transmission, together with via larger respiratory droplets, and direct contact with contaminated surfaces (5,6). Previously, other coronaviruses, (e.g. SARS and MERS) have been shown to disperse via aerosols, and have been determined to cause nosocomial infections as well as extensive hospital outbreaks (7–9). Even if the relative contributions of the different transmission modes remains unclear, the current evidence is sufficiently clear to justify engineering controls targeting airborne transmission as part of an overall strategy to minimize infection risk indoors. Doubtless, this need is particularly important in hospitals and other healthcare facilities managing COVID-19 patients.

Many recent literature studies reported the detection of SARS-CoV-2 in hospitals air samples (10–13). Among these works, a study performed air sampling in the general hospital ward, and detected SARS-CoV-2 PCR-positive particles of sizes >4 µm and 1–4 µm in some rooms, despite these rooms having 12 air changes per hour (12). Scientists have demonstrated that speaking and coughing produce a mixture of both droplets and aerosols in a range of sizes, that these secretions can travel together for up to 4-8 meters, that it is feasible for SARS-CoV-2 to remain suspended in the air and viable for hours, that SARS-CoV-2 RNA can be recovered from air samples in hospitals, and that poor ventilation prolongs the amount of time that aerosols remain airborne (10, 14).

For this reason, in order to improve indoor air quality and reduce the risk of transmission, the World Health Organisation (WHO) recommended a minimum ventilation rate of 288 m^3^ per person (15). However, even where the ventilation systems run at optimal rate there are still frequent cases of airborne bacteria and viruses causing hospital infections. To contain the spread of the virus and reduce transmission and infection rates, air purifiers can act as a supplementary measure (16). Studies show that air purifiers in dental clinics have reduced the transmission of airborne pathogens and exposure to health workers via droplets and aerosols (17). Unfortunately, most air purifiers that are capturing air pollutants by using ventilation, seem not to be completely effective. In a 2003 work, five experiments were conducted to assess how aerosolized bacteria and spores respond like particulate contaminants to the primary electrical forces in a room (18). Most are too small to respond to gravity and ventilation, thus remaining suspended in the air causing potential contagion. This concept can help to better understand viruses’ possible spreading dynamics.

In this work the AIRcels, air purifiers manufactured by a team of researchers at U-Earth Biotech Ltd, were used. The AIRcel attracts the pollutants through a concentration gradient and not through ventilation, then captures them into a water tank where charged particles are removed by electrical grounding and the organic compounds are oxidized. This technology has undergone intensive field testing in hospitals (surgery rooms, emergency rooms and labs), waste treatment plants (19), and heavy-duty manufacturing environments (20, 21) since 2011, proving itself excellent at capturing and destroying a very broad range of contaminants.

Currently, the reference method for the detection of SARS-CoV-2 is the reverse transcription polymerase chain reaction (RT-PCR). Although RT-PCR has high sensitivity and specificity, it requires skilled personnel, and a long period of data processing and analysis (22). Several types of SARS-CoV-2 real-time RT-PCR kit have been developed and approved (23, 24). In this work the multiplex platform (XABT) Multiple Real-Time PCR Rotor-Gene was used. For qualitative immediate detection of SARS-CoV-2, the rapid antigen tests are also available (25). The RT-PCR test is conducted using different types of specimens, including sputum, nasopharyngeal swabs, pharyngeal swabs and saliva. Both RT-PCR and rapid antigen tests are principally performed on symptomatic patients and on those which were in close contact with positive patients. However, many virus carriers are asymptomatic and without the ability to screen them quickly and effectively, they can have the potential to increase the risk of disease transmission if no early effective measures are implemented (26, 27). Therefore there is an urgent need for non-invasive systems that can be used to localize the source of the infection and act as an early warning.

In this regard, a research article illustrates the wastewater-based epidemiology (WBE) analysis, as an effective approach to predict the potential spread of the infection by testing for infectious agents in wastewater, since faeces and urine from disease carriers in the community contain many biomarkers (28). Wastewaters collected in epidemic circulation areas like Milan and Rome were found to be positive for the SARS-CoV-2 virus, according to clinical data, RNA was detected with 6 samples out of 12 testing positive (29).

In the first part of this article some unpublished results from a sampling campaign at Saronno hospital in 2011 are presented. These results show the AIRcel efficiency at capturing and destroying particulate matter and airborne pathogenic bacteria. In the second part of the article we tested the AIRcel bioreactor efficiency on detecting, capturing, and destructing the SARS-CoV-2 virus. The AIRcel units were placed in three different hospitals in Milan, Italy during the COVD-19 health emergency. Some water samples from the AIRcels were collected and sent to the laboratory to be tested for SARS-CoV-2 through real-time RT-PCR. Finally, we developed U-Alert, a protocol to achieve early detection of SARS-CoV-2 virus within the environment via rapid biomonitoring testing through rapid antigen test on the AIRcel water. Given the very promising results we recommend health organizations and local governments to install such protective measures to contain the spread of the virus in the interest of the people.

## MATERIAL AND METHODS

### U-Earth Biotech Ltd. ‘AIRcel’ Technology

‘AIRcel’ is a biological air treatment device manufactured by a team of researchers at U-Earth Biotech Ltd, a biotech company located in the UK and Italy, specializing in biological air purification in work environments. AIRcel is a bioreactor with a smart operating system that captures macro and micro molecules by ionic differentiation and, with the help of the active bio-oxidation process inside, recirculates ‘purified’ air outside the reactor. The unit is replenished weekly with water (which condenses inside the unit) and with a monthly dose of U-Ox additive (non-pathogenic, non-GMO), which is a consortium of microorganisms that digest air contaminants. These bioreactors, in correlation with a biofilter technology, consist of various phases in close contact such as a solid, liquid, and gas phase. The bioreactor itself is considered the solid phase, the liquid phase consists of water, and treated air is considered the gas phase. The usual biofilter design consists of physical support for biomass, whereas the patented plastic bioreactor consists of an optimized configuration in order to enhance the biomass-degrading activity, ‘digesting’ captured contaminants by bio-oxidation. Pollutants are attracted and captured via liquid-gas mixing through the reservoir tank which contains electrically grounded water that cleans and purifies the air. In these bioreactors, charged particles attract the contaminants and any odors generated, and they are removed through electrical grounding wherein organic compounds are oxidized (30, 31). Generally, pollutants move from high concentration to low concentration enabling the process to repeat itself as a sustainable technology wherein air quality is maintained. Unlike other air treatment systems, it does not require pressure membrane filters or high temperatures to operate. By attracting airborne pathogens carried by aerosols in rooms, which ordinarily deposit on surfaces, it helps maintain hygienic ventilation.

### Instrumentation during the Saronno hospital campaign in 2011

The campaign at Saronno hospital in 2011 included the installation of one AIRcel 600 and four AIRcel 85 systems (two models of air purifiers that only differ from dimensions and capacities) into a highly frequented hospital area of 1000 sqm with a footfall of around 1300 people per day. The units were placed in visiting rooms, the central booking office, and a waiting area for 100 people. To verify the effectiveness of the system, monitoring activity on indoor air quality was performed for a one-month duration (from 02/08/2011 to 05/09/2011). The monitoring activity was also performed on processed water quality to evaluate the fate of contaminants captured. The trial was carried out in normal operating hours, with no sealing of the indoor environment (door and windows open or closed-depending only on staff/patients’ needs).

For particles’ sampling the AEROTRAK™ Portable Airborne Particle Counter (ISO 21501-4:2007) was used for cleanroom particles classification following ISO 14644-1:1999 (0.3; 0.5; 1; 2; 3; 5 μm). For microbiological air sampling, the SAS Super

IAQ Surface Air System (model 90593), which conveys a known volume of air during a fixed period on Petri Plates filled with Standard Plate Count Agar (PCA) was used. Samples were then cultivated and the colonies formed were counted after characteristic intervals. Microbiological analysis and standard chemical analysis (including Fluorides, Chlorides, Ca, Mg, K, Cu, Fe, Mn, As, total organic carbon and conductivity analysis) were performed on water samples collected from the AIRcels water.

### Instrumentation during the SARS-CoV-2 pandemic 2020

Ten AIRcel units per hospital were placed in three different hospitals in Milan, Italy during the COVID-19 health emergencies. At Sesto San Giovanni (Multimedica) hospital (Figure 1a) the AIRcels were placed in the COVID-19 dialysis and visiting rooms. At San Raffaele hospital (Figure 1b - note: the air extraction was present on the wall behind every bed), the units were placed in the emergency room (ER) and in the COVID Intensive Care dedicated ward, used for the most serious cases, equipped with all the proper ventilation requirements (contamination most likely occurred during the application of ventilators on intubated patients). AIRcel bioreactors were also placed in other COVID-19 treatment areas including San Giuseppe hospital (Figure 1c) in the obstetrician, ER, and canteen areas.

**Figure 1:**
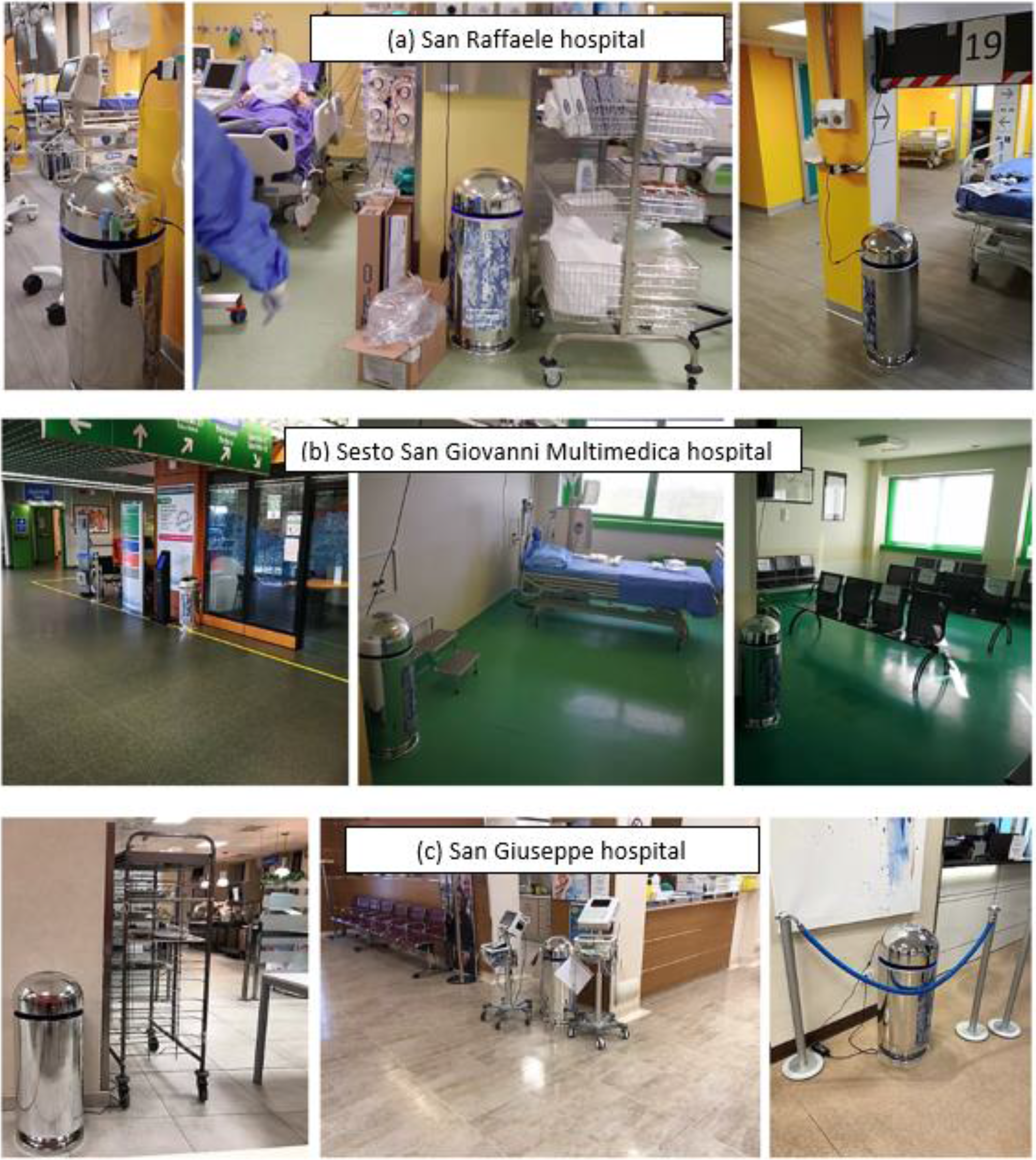
AIRcels setting at San Raffaele hospital (a), Sesto San Giovanni (Multimedica) hospital (b) and San Giuseppe hospital (c) in Milan, during the COVID-19 health emergency.

### The multiple real-time PCR kit

Periodically, some water samples were extracted from the AIRcels and sent to our laboratory for SARS-CoV-2 detection based upon conventional RT-PCR analysis. RT-PCR analysis involves radioactive isotope markers to detect genetic materials of the SARS-CoV-2 pathogen. The analysis is based upon biomolecular assay and sensitive for mRNA detection. The target sequences have been identified and made public since December 2019, therefore the manufacturers of the detection kits were able to market products useful for identifying the target sequence responsible for the pandemic in the early months of 2020.

For the real-time PCR we used a detection system called (XABT) Multiple Real-Time PCR kit for detection of 2019-nCov adapted by our research laboratory in Bolzano by Beijing Applied Biological Technologies Co. Ltd. The kit is based on a multiplex platform capable of simultaneously detecting an extended group of β-coronaviruses. This commercial kit consists of two distinct master mixes identified by tube A and tube B. The first is specific to the target SARS-CoV-2 gene ORF1ab + N, while the second contains the generic β-coronavirus E gene. We tested the kit with extremely diversified types of samples: in addition to the classic buccal, oropharyngeal samples and faecal samples, solute samples were extracted from AIRcel bioreactors. To test the solutes (water + U-Ox additive) from the AIRcels, the QIAGEN’s Pathogen extraction kit supplied by QIAGEN, (Hilden Germany) was used.

### SARS-CoV-2 antigen biological sampling tests

The AIRcel water samples were also tested with the rapid antigen tests. The SARS-CoV-2 rapid test cassette antigen (nasopharyngeal swab) is a rapid qualitative membrane-based immunoassay for the detection of SARS-CoV-2 antigens typically in human nasopharyngeal swab samples. In this study we showed a variant of this test used on environmental samples on a water basis. The SARS-CoV-2 antibody coated region of the test line, when in contact with the sample, reacts with the test SARS-CoV-2 antibody coated particles. The mixture then migrates upward on the membrane by capillary action and reacts with the SARS-CoV-2 antibody in the test line region. If the sample contains SARS-CoV-2 antigens, a colored line will appear in the test line region as a result of the reaction. Positive results indicate the presence of viral antigens, but a verification with a molecular method in the laboratory with PCR protocol is necessary for definitive confirmation. From the tests carried out on solute samples (water + U-Ox additive) taken from the AIRcels it is possible to verify in only 15 minutes whether the device has captured and incorporated suspended airborne particle ‘droplets’ contaminated by SARS-CoV-2, circulating within the range of action of the device itself.

## RESULTS AND DISCUSSION

### Background: Technology testing history on airborne contaminants at Saronno Hospital

The objective of this study, performed in 2011, was to implement the first application in Europe, to achieve the Italian Health and Environment Authority validation and also to verify the system efficiency in a highly frequented hospital environment. This study involved U-Earth, as the system provider, ASL (Health Authority) as hospital counterpart, and ARPA Lombardia (Environment Authority) for monitoring activities and data validation. These results are presented to better understand the capture and destruction dynamics of the technology that occurred during the SARS-CoV-2 testing in Milan hospitals during the pandemic since, during the COVID-19 crisis in hospital wards, was not possible to test all contaminants for safety reasons. It is important to mention that despite the already remarkable results since 2011, both the bioreactors and the biomass additive formula have undergone major improvements to better address the capture/decomposition process.

The main results of the campaign are shown in Figure 2 and Table 1. On the left side of the figure the abatement of bacteria counts (CFU) concentration over the month of the campaign monitoring is shown (also shown in Figure 3). Overall, in the waiting room there is an average reduction of 57% in the CFU concentration with a maximum reduction of 87% (Figure 2a, Table 1); an average reduction of 87% and a maximum reduction of 98% in the central booking office (Figure 2b, Table 1), and an average reduction of 87% and a maximum reduction of 95% in the visiting rooms (Figure 2c, Table 1). On the right side of the figure the abatement in the particle number concentration in the visiting rooms per different particle ranges is shown.

**Table 1:**
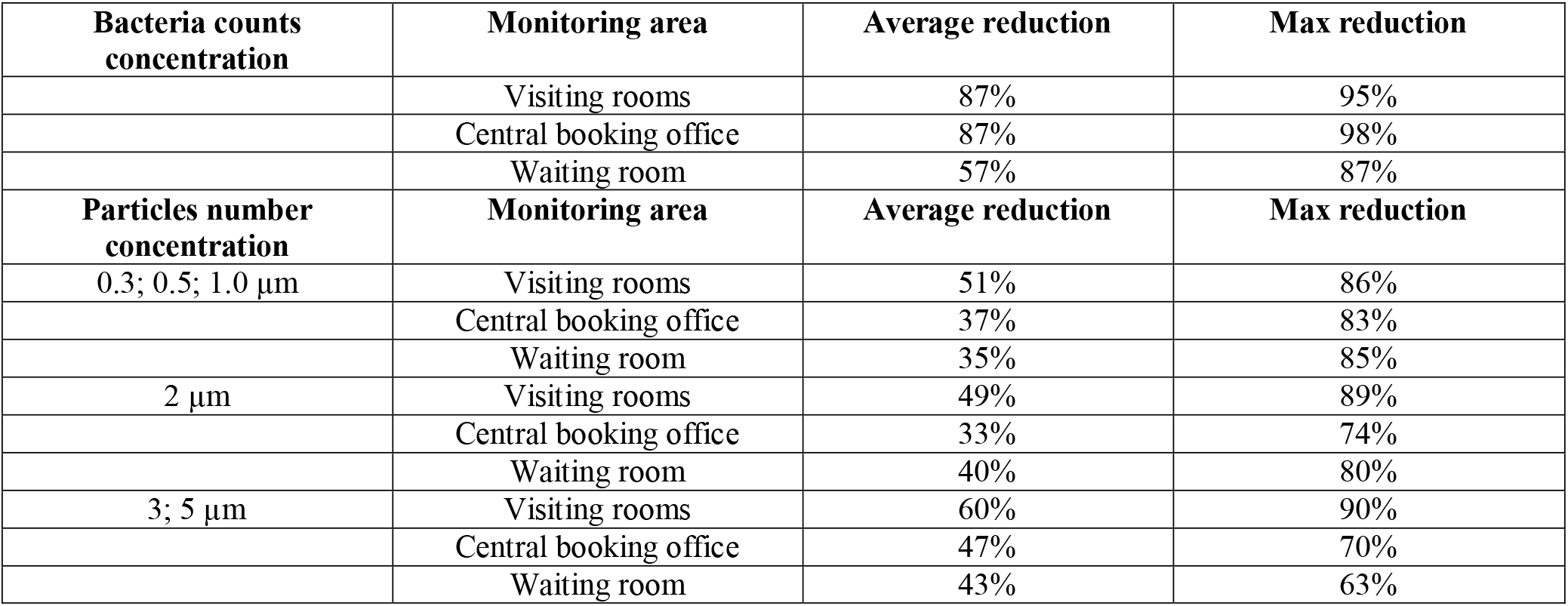
Average and maximum (max) reduction per cubic meter per bacteria counts and particle number per different size ranges.

**Figure 2:**
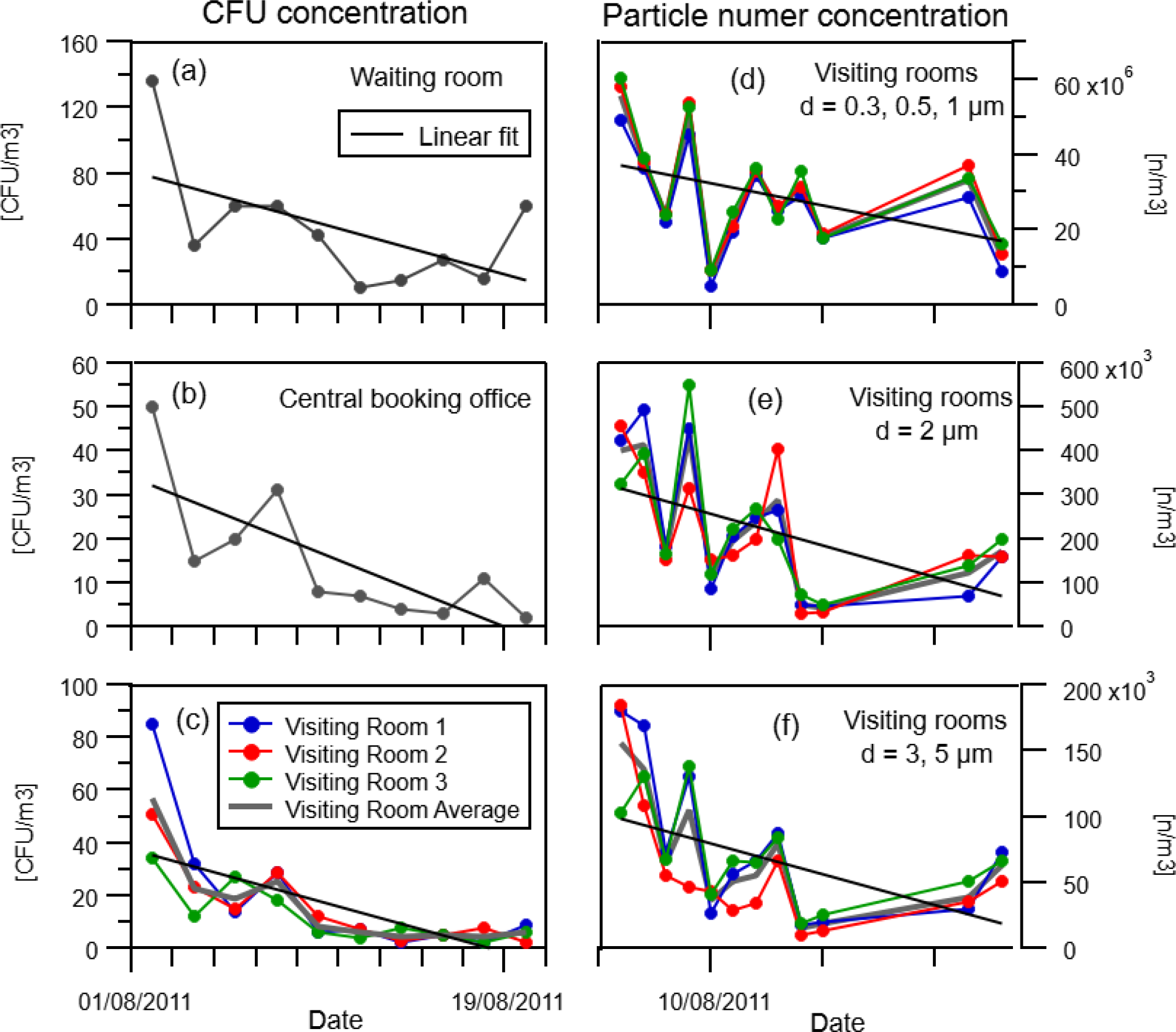
The left side of the figure shows the abatement in CFU concentration in the waiting room (a), central booking office(b) and visiting rooms (c). The right side of the graph illustrates the abatement in particle number concentration in the visiting rooms per different particle size ranges: 0.3, 0.5 and 1 µm (d); 2 µm (e) and 3, 5 µm (f).

**Figure 3:**
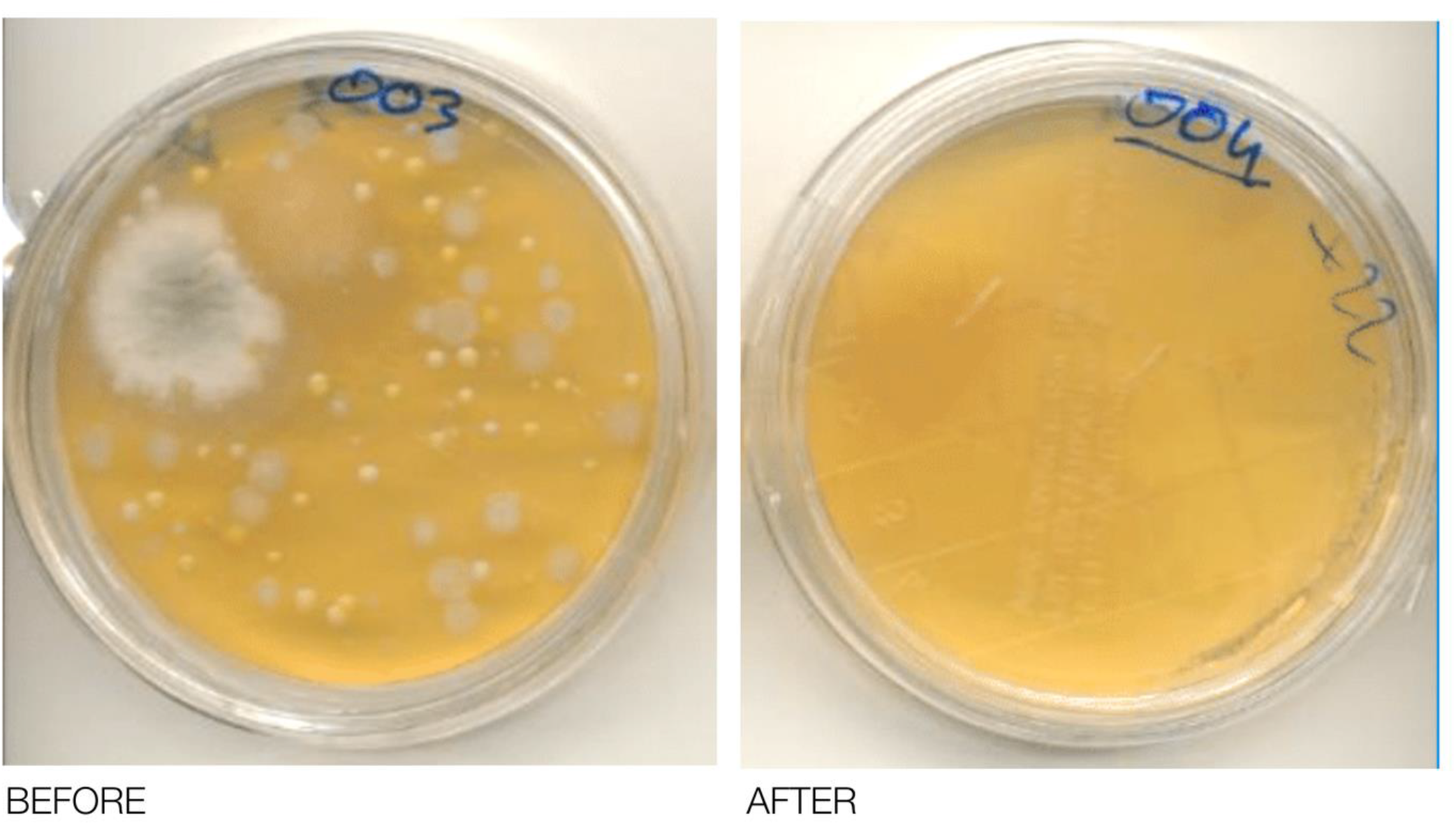
An example of some Petri Plates filled with Standard Plate Count Agar before and after the AIRcel placement.

As shown in Table 1, remarkable reductions in the number concentration of particles, from an average value of 37% to 69% was obtained per all the size classes. It is important to point out that this abatement trend was obtained in normal operating hospital activities, while usually the air purifiers tests are run in a laboratory that do not fully represent real case scenarios.

Pathogenic bacteria such as Legionella, Enterovirus, Escherichia Coli, Enterobacteria, Enterococcus, and Salmonella counts were checked in the processed water of the, AIRcel after two and three months of system activity (05/10/2011 and 08/11/2011), and the results were compared with tap water supplied in the hospital. The aim was that of verifying possible changes in the qualitative characteristics of the AIRcel water and if potential changes could be related with the activity of AIRcel in capturing the aero disperse pollutants. The findings showed Staphylococcus and Pseudomonas presence (in lower concentration that the maximum values referrable to drinkable water) in the AIRcel water sample and not in the tap water of the hospital. No bacteria presence was found in the second water sample on 08/11/2011. The results were interpreted by all the institutions involved, as a capture of the aero disperse Staphylococcus and Pseudomonas bacteria by the AIRcel and their successive digestion.

### During the COVID-19 crisis: Technology Testing for SARS-CoV-2 at Multimedica, San Raffaele and Sacco hospital

In total 68 samples were processed in three distinct test sessions between April and June 2020, using the QIAGEN Rotor-Gene thermal cycler. The result of the RT-PCR showed a marked presence of the target β-coronavirus E gene for 19 of the 68 samples, while the target ORF1ab + N was detected in 7 samples. In particular, at Sacco Hospital, the test results show the detection of ORF1ab + N and/or E gene in 15 samples out of 40.

Further experimental activity and related tests were conducted on the acquired samples. Twelve water samples of 100 ml each were selected and collected from hospital units that tested positive for both viral targets, and they were treated with 1 ml of U-Ox microbial additives on an equal volume of sample to test the short-term degradation capacity in vitro. In 5 out of 12 samples, traces of the viruses were no longer found after the overnight period. These results indicate the system’s ability to capture virus droplets and destroy them inside the reactor. Based on these results we interpreted the results shown in Table 2 in the same way : in many samples SARS-CoV-2 virus was first captured and then digested by the AIRcel.

**Table 2:** Test results show in Multimedica hospital the detection of ORF1ab + N and/or E gene in 4 samples (a); test results show in San Raffaele hospital the detection of ORFlab + N and/or E gene in all 6 samples (b); test results show in Sacco hospital the detection of ORF1ab + N and/or E gene in 15 samples out of 40 (c).

| (a) | Multimedita hospital |  |  |  |
| --- | --- | --- | --- | --- |
|  | 30/04/2020 | 29/05/2020 | 04/06/2020 | Notes |
| Sample 1 |  |  |  |  |
| SARS-CoV-2 | Detected | Digested | Not Detected | The water was not changed b/w the 2 testing rounds |
| β-coronavirus E gene | Detected | Digested | Detected | The water was not changed b/w the 2 testing rounds |
| Sample 2 |  |  |  |  |
| SARS CoV-2 | Not Detected | Not Detected | Not Detected | - |
| β-coronavirus E gene | Detected | Digested | Not Detected | The water was not changed b/w the 2 testing rounds |
| (b) | San Raffaele hospital |  |  |  |
|  | 30/04/2020 | 29/05/2020 |  | Notes |
| Sample 1 |  |  |  |  |
| SARS CoV-2 | Not Detected | Detected |  | - |
| β-coronavirus E gene | Not Detected | Detected |  | - |
| Sample 2 |  |  |  |  |
| SARS CoV-2 | Not Detected | Detected |  | - |
| β-coronavirus E gene | Detected | Digested |  | The water was not changed b/w the 2 testing rounds |
| Sample 3 |  |  |  |  |
| SARS CoV-2 | Not Detected | Not Detected |  | - |
| β-coronavirus E gene | Detected | Detected |  | - |
| Sample 4 |  |  |  |  |
| SARS CoV-2 | Not Detected | Not Detected |  | - |
| β-coronavirus E gene | Not Detected | Not Detected |  | - |
| (c) | Sacco hospital |  |  |  |
|  | 04/06/2020 | 17/08/2020 |  | Notes |
| Sample 1 |  |  |  |  |
| SARS CoV-2 | Not Detected | Not Detected |  | - |
| β-coronavirus E gene | Detected | Detected |  | - |
| Sample 2 |  |  |  |  |
| SARS CoV-2 | Not Detected | Not Detected |  | - |
| β-coronavirus E gene | Not Detected | Detected |  | - |
| Sample 3 |  |  |  |  |
| SARS CoV-2 | Not Detected | Detected |  | - |
| β-coronavirus E gene | Not Detected | Detected |  | - |
| Sample 4 |  |  |  |  |
| SARS CoV-2 | Detected | Digested |  | The water was not changed b/w the 2 testing rounds |
| β-coronavirus E gene | Detected | Digested |  | The water was not changed b/w the 2 testing rounds |
| Sample 5 |  |  |  |  |
| SARS CoV-2 | Not Detected | Detected |  | - |
| β-coronavirus E | Not Detected | Not Detected |  | - |
| **gene** |  |  |  |  |
| **Sample 6** |  |  |  |  |
| **SARS CoV-2** | Not Detected | Not Detected |  | - |
| **β-coronavirus E gene** | Not Detected | **Detected** |  | - |
| **Sample 7** |  |  |  |  |
| **SARS CoV-2** | Not Detected | **Detected** |  | - |
| **β-coronavirus E gene** | Not Detected | **Detected** |  | - |
| **Sample 8** |  |  |  |  |
| **SARS CoV-2** | Not Detected | Not Detected |  | - |
| **β-coronavirus E gene** | **Detected** | **Detected** |  | - |
| **Sample 9** |  |  |  |  |
| **SARS CoV-2** | Not Detected | Not Detected |  | - |
| **β-coronavirus E gene** | Not Detected | **Detected** |  | - |
| **Sample 10** |  |  |  |  |
| **SARS CoV-2** | Not Detected | Not Detected |  | - |
| **β-coronavirus E gene** | Not Detected | **Detected** |  | - |

This grants AIRcels full-fledged inclusion as a valid support for biomonitoring of the areas where they are installed. AIRcels were found to be effective in hospital wards where COVID-19 positive patients or suspected asymptomatic patients were kept for observation. The air purifiers reduced the viral load by capturing and destroying the SARS-CoV-2 virus in their vicinity and therefore reduced exposure to the virus.

Another session of experiments was conducted to test a new rapid diagnostic kit for the detection of SARS-CoV-2 antigen, on samples that already tested positive for the specific target when processed with the real-time PCR-RT. The purpose of the experiment was to correlate the same methods used to detect the SARS-CoV-2 target for human use to a new environmental level, in order to have a useful detection system for the AIRcel users to run on their own and get results in less than 15 minutes. Both positive and negative samples were then compared to verify the sensitivity of the rapid kit and its use, since the CE IVD validation required its use for diagnostic purposes only. The same quantity as foreseen by the protocol, validated with a nasopharyngeal swab, was tested by directly entering the solute extracted from the AIRcel water tank into the test panel cassette of the chromatographic support used to perform the test. All the samples gave the same results as the RT-PCR. These very promising results allowed us to develop a protocol we called U-Alert for continuous environmental biomonitoring and quick virus detection of SARS-CoV-2 RNA through analysis of the recirculating water of the AIRcel, first with the rapid antigen diagnostic kit, then by confirmation of the results via RT-PCR. In this way, the complete virus detection protocol provides early warning that quickly confirms (within 24 hours) the presence of the viral targets being detected.

## CONCLUSION

This study has demonstrated the capability of AIRcel bioreactors to capture and digest viruses such as SARS-CoV-2 and β-coronaviruses. It is also important to note that it is possible to carry out this checking activity periodically, on a scheduled basis, for the placed AIRcels. The continuous monitoring activities can provide useful data on the biological activity of bioreactors over time, to evaluate how the microbial community present in the devices interacts with the virus. In this study, AIRcel bioreactors have demonstrated an analytical approach to maintain air quality and quantify the presence of viral targets through efficient biomonitoring leading to the capture and destruction of SARS-CoV-2 and β-coronaviruses. The use of AIRcels as an environmental monitoring tool, especially if combined with the rapid detection system of the SARS-CoV-2 antigen supplied with the air purifier, provides valid support to the user, by giving the possibility of carrying out a preventive diagnosis not only on air quality but also of the actual presence of the coronavirus (with U-Alert protocol), which will then be eventually confirmed with the RT-PCR technique. Detecting the presence of inactivated SARS-Cov-2 in the AIRcels’ water tanks, for example in a school, while lowering the viral charge suspended in the air, could also effectively inform on the possible presence of positive individuals in selected classrooms that require further testing. Accurate detection is crucial for prevention.

## Data Availability

The authors confirm that the data supporting the findings of this study are available within the article.

## Acknowledgment

The authors wish to thank specific individuals who assisted in this study along with hosting the AIRcel units in COVID affected areas, providing samples and assistance crucial for the study: Ospedale San Raffaele team and specifically Dr. Elena Bottinelli and Dr. Matteo Moro, Ospedale Multimedica: Dr. Matilde Ardizzi, Eng. Varisco Dr. Andrea Vergani and Dr. Davide Pasetti. Ospedale Sacco: Dr. Matteo Letzger, Dr. Pietro Olivieri, and Dr. Gabriele Luongo. Special thanks to Betta Maggio and the U-Earth Team for providing background information on the technology, AIRcel equipment, and inspiration for this study.

## Author’s Contributions

All authors critically revised the article and approved the final version for submission for publication.

